# Emergency Department Patients’ Perspectives on Being Offered HIV Pre-Exposure Prophylaxis (PrEP) Services in an Urban ED

**DOI:** 10.1101/2025.02.07.25321883

**Authors:** Rachel E Solnick, Tatiana Gonzalez-Argoti, Laurie J. Bauman, Christine Tagliaferri Rael, Joanne E. Mantell, Yvonne Calderon, Ethan Cowan, Susie Hoffman

**Author notes:** Corresponding Author: Rachel E Solnick, MD, MSc, Icahn School of Medicine at Mount Sinai Hospital Department of Emergency Medicine Research Division 150 E 42nd St New York, NY 10017.

## Abstract

HIV pre-exposure prophylaxis (PrEP) is underutilized in the United States. Emergency Departments (EDs) can be strategic locations for initiating PrEP; however, knowledge concerning patients’ receptivity to ED PrEP programs is limited. This study explores ED patients’ perspectives on PrEP service delivery and their preferences for implementation. Semi-structured qualitative interviews were conducted with 15 potentially PrEP-eligible ED patients to examine their receptiveness to PrEP services, preferences for delivery methods, and logistical considerations. Most participants were open to learning about PrEP in the ED, provided it did not delay care, occur during distress, or compromise privacy. Universal PrEP education was viewed as reducing stigma and increasing awareness, while targeted screening was seen as efficient. Participants strongly preferred receiving information in person rather than via videos or pamphlets. Concerns included ensuring ED staff expertise and maintaining privacy during PrEP-related discussions. Regarding same-day PrEP versus prescriptions or referrals, opinions varied, with participants valuing flexibility and linkage to care. This first qualitative study of ED patients’ perspectives on PrEP services highlights general receptiveness, with key concerns about privacy, expertise, and wait times. Patient-centered approaches, including integrating PrEP services into ED workflows, offering flexible initiation options, and providing privacy, can support the feasibility of ED-based PrEP programs.

## INTRODUCTION

HIV pre-exposure prophylaxis (PrEP) is a highly effective method for reducing HIV transmission.^1–4^ Recent modeling suggests that providing PrEP to populations at higher risk of HIV could reduce new diagnoses by 18%.^5^ Despite its proven efficacy, PrEP uptake in the US remains low due to structural, provider, and individual-level barriers.^6^ Additionally, significant disparities exist by race, gender, and geographic location in PrEP access and delivery^7–9^ and have led to implementation efforts in diverse medical and non-traditional settings.^10,11^

Emergency Departments (EDs) are strategically positioned to reach populations who are disproportionately affected by HIV. EDs serve many underserved, uninsured, or underinsured individuals, including racial/ethnic and sexual/gender minority groups—populations disproportionately affected by HIV.^12,13^ For some, EDs may be their only interface with the healthcare system, suggesting EDs could play a role in PrEP screening, initiation, and referral.^14–19^

Despite its potential, research on ED-based PrEP programs remains limited.^20–24^ Previous research has assessed provider- and setting-related barriers, such as low PrEP awareness among ED clinicians, a focus on acute care, and logistical challenges, such as staffing, financing, and unclear follow-up pathways.^25,26^ Less is known about patient preferences regarding the delivery of PrEP in the ED.^21^ Thus, the objective of this study was to qualitatively examine patients’ preferences across the PrEP continuum of screening, education, initiation, and linkage and how they could be implemented in the ED setting.

## METHODS

### Study design and setting

As part of a study to identify approaches for implementing PrEP services in the ED, semi-structured interviews were conducted with 15 non-acute, potentially PrEP-eligible patients presenting to the ED of Mount Sinai Beth Israel (MSBI) in 2022. Built in 2010 in New York City, MSBI is a 700-bed hospital with an 85-bed ED managing 75,000 patient visits per year with an admission rate of 25%. The racial and ethnic makeup of the hospital population is predominantly Hispanic (51%) and Black (39%). The 2016 payor mix was 38% Medicaid, 27% Medicare, 25% private insurance, and 10% self-pay.

### Participant eligibility and recruitment

Eligible participants were aged 18 years or older, self-reported HIV-negative, English-speaking, and purposive sampled as potentially eligible for PrEP, based on the US Centers for Disease Control (CDC) 2021 guidance criteria^27^ and other recommendations relevant for women,^28^ as shown in **Box 1**. Potential participants were presented with these criteria by a research assistant (RA) and indicated if any applied to them; they did not have to specify which applied. Exclusion criteria included currently taking PrEP, being unwilling to be audio-recorded for the interview, and not having contact information to schedule the interview following the ED visit.

To recruit the sample, RA monitored the health information system ED track board to identify adult patients with complaints related to sexually transmitted infections (STIs), non-occupational post-exposure prophylaxis (nPEP), or injection-related complications. Before approaching the patient, they obtained permission from the patient’s ED provider and confirmed with the provider that the patient was cognitively intact and medically and psychiatrically stable. Potential participants were informed about the research purpose and verbally consented to complete the eligibility screen administered by the RA using REDCap, a HIPAA-compliant data capture system.^29^ Eligible patients were invited to participate in a one-time interview with a study team member designed to take 30-45 minutes.

#### Box 1.

##### Eligibility criteria

###### Does one or more of these apply to you?

- Have had sex or shared needles with someone in the past 12 months who has HIV or whose HIV status I did not know.
- Have been diagnosed with syphilis, chlamydia, or gonorrhea in the past 12 months
- Have taken non-occupational HIV post-exposure prophylaxis (PEP) in the past 12 months
- Have had sex with someone in exchange for money, drugs or housing in the past 12 months
- Have experienced forced sex in the past 12 months
- Think PrEP could be beneficial to me for some other reason

*Note:* The above criteria are based on published guidance^27,28^

### Study procedures

Interviews were scheduled at a convenient time for participants following their discharge from the ED. Three experienced qualitative interviewers (TGA, SH, CTR) conducted the interviews using a HIPAA-compliant virtual platform (Zoom Video Communications, Inc. Version: 5.11.0) and obtained verbal consent. Video files were deleted after the interview, and audio recordings were securely transmitted for professional transcription. Participants were compensated $50 for their time. The study was approved by the Institutional Review Boards (IRBs) of the Albert Einstein College of Medicine-Montefiore Medical Center (IRB #2021-13676), the Mount Sinai Health System (STUDY-21-01811), and the New York State Psychiatric Institute/Columbia University Department of Psychiatry (IRB #8239).

### Interview guide

The interview guide was designed to elicit participants’ responses to receiving PrEP services in the ED and their thoughts about when and how these services should be offered. The guide was based on an ED-PrEP cascade developed in partnership with a Community Collaborative comprising ED physicians and administrators, health department HIV prevention experts, and leadership of community-based organizations (CBO) engaged in HIV prevention (**Figure 1**).

**Figure 1.**
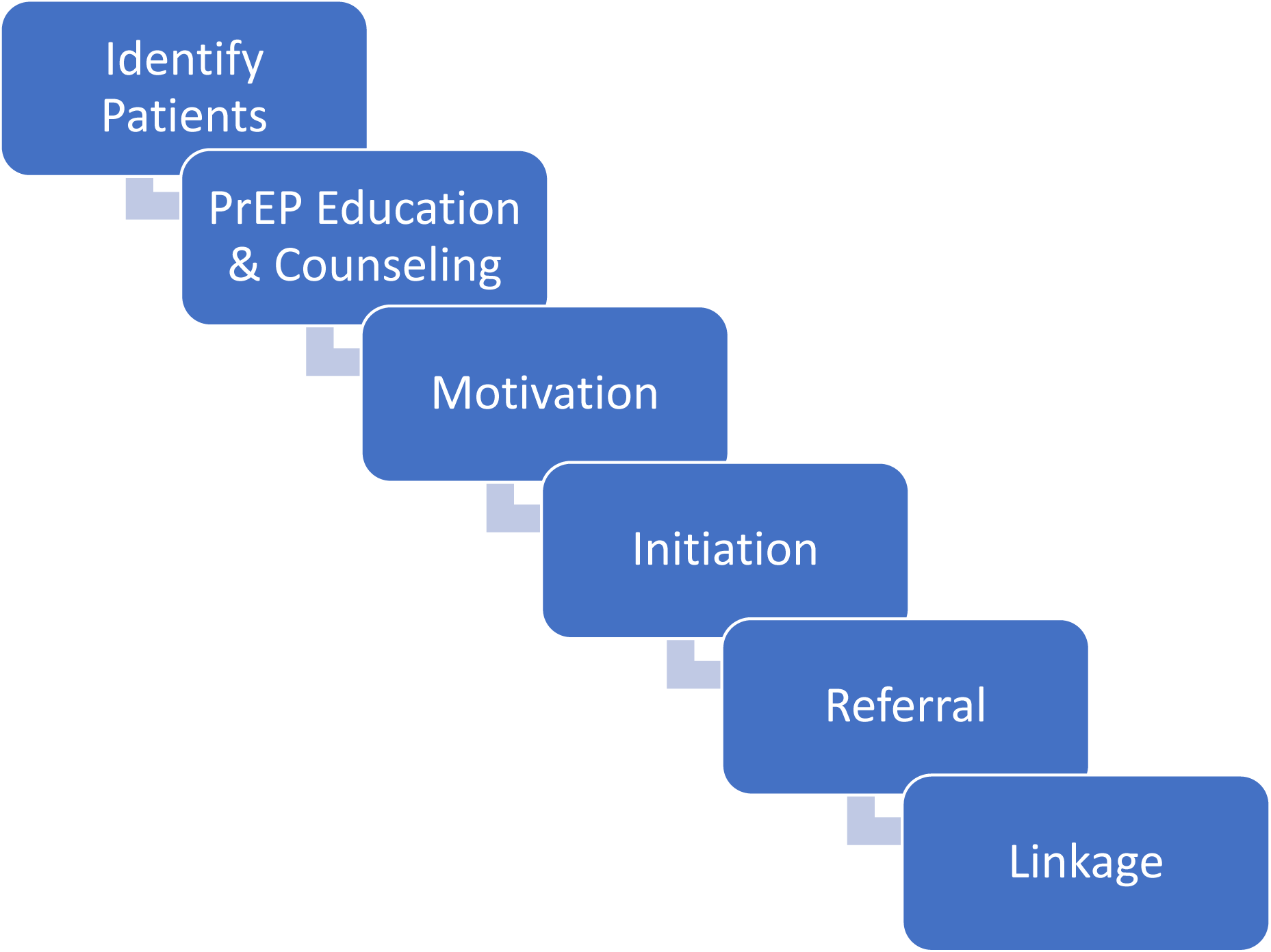
ED-PrEP Cascade

The interview queried patients about their preferences regarding (1) whether PrEP information should be given to all or only specific individuals based on screening; (2) the best time for presenting PrEP information and/or performing screening; (3) who should deliver the information and conduct screening (clinical staff, health educators, or peers); (4) preferences for the mode of education (video on laptop/tablet, pamphlets, or in-person; (5) how much additional time they would be willing to spend in the ED for education or screening; (6) preferences for starting PrEP immediately in the ED versus receiving a prescription for pharmacy pick-up or a referral to another care site; (7) willingness to undergo an additional blood draw for PrEP-related screening; (8) preferences for location of follow-up care – with a primary care provider or a medical site with PrEP experts.

At the start of the interview, participants were given a brief explanation of PrEP and asked how they would respond to being offered PrEP services in the ED. At the end of the interview, participants were asked to reflect on their overall thoughts about receiving PrEP services in the ED.

### Data analysis

Qualitative analysis was conducted using a rapid analysis technique, selected as a methodology that can produce timely findings while maintaining rigor.^30^ We began with a deductive approach, applying broad predetermined codes based on the interview guide topics. Codes were applied to the relevant text using Dedoose (version 9.0.17),^31^ and a coding report was generated for each code. Members of the analysis team were assigned to review and summarize a set of codes, identifying subcodes (e.g., preferences around ways to receive PrEP education in the ED) and any new themes. At regular check-in meetings during the analysis phase, the team discussed and achieved consensus on new themes that emerged inductively. For the final analysis, the first author read all the coding reports and summaries and integrated them into a framework of three key domains for ED-PrEP planning and implementation: (1) <u>Patient characteristics</u> (e.g., perceived risk for HIV, receptiveness to both HIV prevention messaging and receiving those services in the ED); (2) <u>Intervention characteristics</u> (e.g., preferences for who provides the services and timing during the visit, the amount and format of information provided); and (3) <u>Contextual/organization factors</u> (e.g., what role the ED plays in the healthcare system.) All coding team members reviewed and concurred on the final analysis.

## RESULTS

### Participants

Out of 175 patients screened, 57 were eligible, 52 agreed to participate, and 15 completed interviews. One interview was not audio-recorded, so the analysis is based on 14 transcripts and one interview summary. Most participants were under 40 years old (n=9) and male (n=11) (**Table 1**). All had insurance, with eight covered by Medicaid. Participants represented a range of races/ethnicities, with the majority identifying as Latino/Hispanic, Black, or mixed. Most participants (n=8) reported only one ED visit in the last 6 months.

**Table 1.**
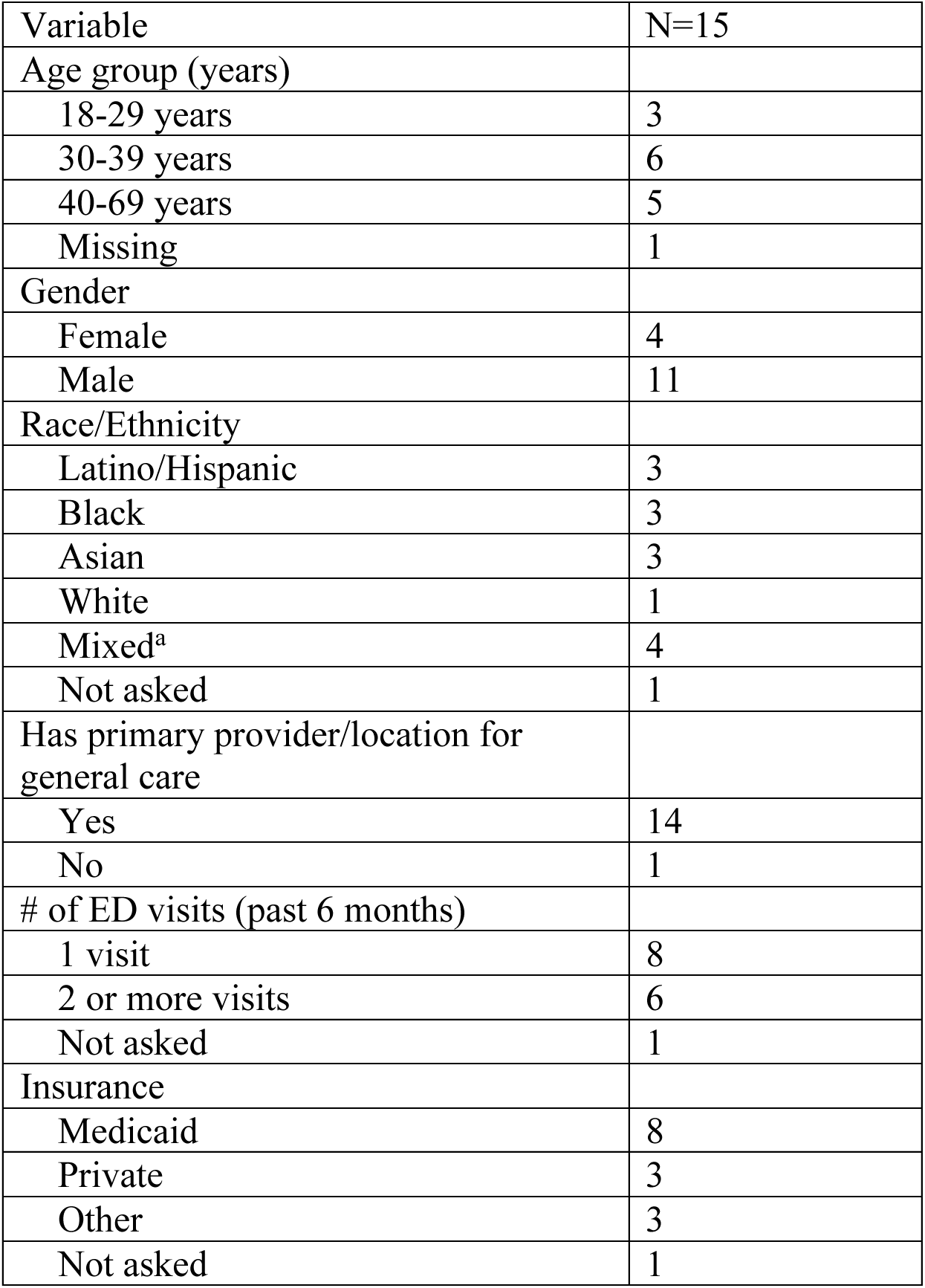
Characteristics of ED Patient Participants.

### Major Themes and Subthemes

**Table 2** displays the major themes, subthemes, and representative quotes described below.

**Table 2.** Major Themes, Subthemes and Representative Quotes.

| **Table 2. Major Themes, Subthemes and Representative Quotes** |  |  |  |
| --- | --- | --- | --- |
| **Domain** | **Theme** | **Summary** | **Representative Quotes** |
| Patient Characteristics | Receptiveness to PrEP | Most participants felt the ED was a setting they were willing to learn about PrEP, however a few were not willing, due to concerns of having more pressing medical issues or that the topic was inappropriate to discuss in the ED. | "I would be interested in hearing about it. It depends on why would be in the ER [ED]. It might be minimal concern at that point but yeah more information is best" (#232)<br>""I don't know if that's something that y'all should be asking or we should be volunteering this information." (#125) |
|  | Perceived Risk for HIV and Motivation | Some participants appreciated that PrEP was more relevant to them based on their behavior. Other participants noted it was a helpful start to a conversation that could be followed up outside the ED. | "I didn't know about PrEP before I came to the emergency room, but I can see how it could help someone like me" (#157). |
| Intervention Characteristics | Preferred Timing and Screening Approach | Most participants preferred screening after registration, emphasizing that it should not interfere with the primary reason for the ED visit. Supporters of universal education highlighted stigma reduction, educating hard-to-reach people, spreading knowledge. Supporters of targeted education emphasized resource efficiency and relevance. | "Once they're stabilized and their immediate needs are taken care of then maybe consider approaching them with the question" (#066). "...educate everyone because of the stigma around the whole virus..." (# 002) |
|  | Education Medium Preferences | A majority preferred personal interaction for receiving PrEP information, valuing engagement. Videos and pamphlets were seen as impersonal, with a few suggesting a hybrid approach incorporating multiple media to suit individual preferences. | " when you speak with someone and directly there is a conversation, there's engagement, there's social cues, there's things that you can look out for. (#232)" |
|  | Preferred Providers for Education | Opinions were split on whether clinicians or non-clinicians should conduct the screenings, with an emphasis on the staff's knowledge rather than their official role | "...like maybe [the peer's] experience is personal and [might] not fit my situation." (#157) |
|  | Initiation and Follow-up Preferences | Preferences for starting PrEP were split between immediate initiation in the ED, receiving a prescription for later, or being referred to a follow-up site. Follow-up preferences were varied, with some preferring their PCP and others a specialist | "Only because of, of that patient-doctor relationship that we already have..." (112) |
| Contextual/<br>Organizational Factors | Benefit of the ED as a General Catchment for Healthcare | Several felt positive towards the ED as it had a catchment area for a potential higher need population, and that there was less stigma around going to the ED compared to a sexual healthcare site. | "Basically, it gives people a chance to, you know, if they're scared to go to a sexual clinic, they have the option to go to a hospital." (#051) |
|  | ED Busyness | Some participants mentioned long ED wait times and a desire for more convenience. A few suggested using this waiting period for activities like PrEP education to improve efficiency and make productive use of their time. | "I was probably waiting for about an hour and a half before I actually received a bed for my care. So, if I had someone, something to do, even if it was like 10, 15 minutes during that waiting period, I would at least feel like I'm working towards being seen or like something medically happening." (#232) |
|  | Time Burden and Convenience | Some participants mentioned long ED wait times and a desire for more convenience. A few suggested using this waiting period for activities like PrEP education to improve efficiency and make productive use of their time. | "I hear this information while I wait and it's not making me lose my spot then I don't see why not." (#232) |
|  | Privacy Concerns | Some emphasized the importance of privacy when discussing PrEP, preferring private spaces or video resources over public areas like waiting rooms to maintain confidentiality. | "Because it definitely wouldn't be confidential because the person is right there." (#164)<br>"that should be done during the nurse's visit. Like when, when you're like in a more private setting because it has a fear of you know, because the fear of everyone else around you." (#051) |

### Key Domain 1: Patient Characteristics Receptiveness to PrEP services in the ED

Most participants expressed interest in and a willingness to learn if they are offered PrEP services in the ED. Several expressed an enthusiastic desire for more medical information, especially for highly effective interventions that they may not have been aware of: “I will absolutely be willing to listen to all of the information… the number is great like you have like 99%” (#157) stated one participant, referring to the reduction in HIV risk. A few participants supported spreading awareness: “A lot of people don’t know about good medicines” (#065) and “I would be 100% [for] receiving the information…” (#058).

A few individuals stipulated their willingness depended on not having an urgent condition and not being in significant physical pain. One participant noted, “If I’m in there for something more life-threatening, it might not be the best time, but if it’s something quick and I hear this information while I wait, then I don’t see why not” (#232). A few participants expressed hesitation about receiving information unrelated to their immediate medical needs but were still willing to receive education, “It depends on why [I] would be in the ER. It might be a minimal concern at that point, but yeah, more information is best” (#232). One person expressed that it was inappropriate to receive PrEP education in the ED: “When you do go to an ED, people are worried about greater things than learning about [PrEP] … there’s like a time and a place for everything” (#192).

### Perceived Risk of HIV and Motivation

Participants expressed greater interest in PrEP education when they perceived it to be relevant to their personal HIV risk. One participant suggested they had had a potential sexual exposure to HIV: “I’m in that situation for the last few days, so I will … be very happy to know” … “I can see how it could help someone like me” (#157).

Even if they did not currently perceive themselves to be at risk for HIV, many participants were still open to receiving the information. One person indicated it might be helpful to know about PrEP for the future: “It might be a minimal concern at that point, but yeah, more information is best” (#232). Others stated, “As someone who is low risk, I would still be interested in learning more” (#088); “I’m not young enough and sexually active enough that I think that I would need that, but I just never know…” (#164).

### Key Domain 2: Intervention Characteristics Preferred Timing of PrEP Education and Screening

There was no consensus on the best time to offer PrEP education or screening, with opinions ranging from during or after triage, before or after seeing a clinician or having tests done, to after the ED visit. The “best time” was seen as situational, depending on each patient’s experience in the ED. Some participants suggested that patients be provided information during triage or while awaiting test results to be engaged without disrupting their care. One participant shared, “Once they’re stabilized and their immediate needs are taken care of, then maybe consider approaching them with the question” (#066). Others preferred receiving information earlier in the visit to avoid prolonging their stay; as noted by one, the drawback of waiting until after the visit was that by then, “you just want to get the hell out of there” (#232).

### Universal vs. Targeted Screening

Participants were evenly divided regarding whether PrEP education should be offered universally or targeted to specific groups based on screening, with some recommending a mixed approach. Those who endorsed universal PrEP education highlighted the benefits of reaching individuals less likely to know about PrEP. Several participants emphasized that the ED could serve as a critical access point for initiating conversations about sexual health, particularly for individuals who might not seek care elsewhere. One participant explained, “It’s a great place to plant the seed…” (#232), possibly leading them to have more conversations with their primary care provider (PCP). A few participants emphasized the importance of destigmatizing PrEP, advocating to “educate everyone because of the stigma around the whole virus…” (#002).

Conversely, a few participants preferred targeting education to those most interested in PrEP or most likely to benefit based on HIV likelihood screening. As one participant explained, “I think someone who takes the time to answer those [screening] questions, they’d be more open to learning about things maybe or actually responding to folks” (#063).

Several participants proposed a combined approach, suggesting that universal PrEP education and screening could be directed to populations more likely to benefit, i.e., targeting people between ages 18-30 years, a period when they are “kind of like not wilding out but experiencing sex”(#039), and another thought it should for individuals in high HIV prevalence areas, “Spanish Harlem…was a high-risk area for children as well as young adults” (#164).

### Education Medium Preferences

More than half of participants preferred receiving PrEP information in the ED primarily through personal contact, which was thought more engaging and easier to understand than education via videos or pamphlets. Some expressed concern that non-interactive methods could lead to disengagement because, otherwise, things might get “lost in translation” (#164) or that one may “zone out […] if someone just handed me a tablet to view” (#232).

Several participants said pamphlets/flyers were not engaging and less effective, leaving individuals feeling “disconnected” (#66); they would likely get lost or thrown out because “we get so many pamphlets” (#232). However, some participants thought printed information had value as a helpful adjunct to personal interaction. It was noted that pamphlets could include links and phone numbers for additional information following the ED visit or that links could be provided electronically via email or QR code for further education.

A few participants preferred learning about PrEP through videos, which might appeal to younger people accustomed to visual platforms: “Younger people grew up in the internet age. We have Instagram, YouTube. We’re more of visual learners, right” (#002). Another said a video could be a “happier and more fun…” distraction, such as TikTok (#157).

### Preferred Providers for PrEP Education

Participants were split on who should deliver PrEP education: clinicians (e.g., nurses or physicians) or non-clinicians (e.g., peer counselors or health educators). Both clinicians and health educators were considered knowledgeable in conducting PrEP screening and education. Some participants did not have strong feelings about the staff’s background, “Doesn’t matter as long as … they know what they’re talking about” (066). Some preferred clinicians: “Everyone’s most trusting of like the doctor that’s taking care of you, because they do have more experience” (#192).

Opinions were divided regarding peers. One participant was worried that peers might not have the necessary competencies, “not sure how much I’d actually think I’d benefit from speaking to only a peer.” Still, when explained by the interviewer that they would be knowledgeable staff, this participant changed their mind, stating, “That makes more sense when I think about it … I would trust that person more, knowing they have similar life situations.” Another was worried a peer was “not like professional” and concerned their lived experience would “not fit into my situation,” stating instead they preferred a health educator because they were “happy to get to know stuff about HIV from someone a similar age as me” (#157).

A few preferred peer counselors because they felt they would be “someone who can relate to the person” (#232). One participant reflected that it depended on the specific task, noting that they would prefer the clinical experience of someone to speak with initially for the “hard questions” and then later for medication administration questions, they could talk with someone “a peer, someone who has been on PrEP or worked with the pill for a while, an educator” (#002).

### Initiation and Follow-up Preferences

Participants had mixed preferences regarding PrEP initiation. Some preferred same-day PrEP in the ED, some opted for a prescription sent to their pharmacy, and some preferred starting at a follow-up site. One participant suggested offering all three options to allow patient choice (#051). Participants who wanted a same-day PrEP preferred the convenience and the peace of mind afforded by immediate initiation: “I would try it right away instead of calling the pharmacy” (#002). This participant added that immediate PrEP initiation in the ED could provide reassurance: “…if you want immediate help, when you have to wait, it can trigger people with doubt, paranoia, worry… if you can get it right away, it’s going to give you a sense of comfort (#002). Another individual said they preferred the same-day start based on a prior experience in which they were concerned about HIV risk. If there were a delay, it would have negatively impacted their health: “I felt right when they gave it to me, right there and then, it was just a sign that people really do care for others” (#058).

Those who preferred a delayed start cited reasons such as having a waiting period to consider questions to ask an expert, having time to reflect, enabling ongoing care for PrEP and other health issues, and ensuring their primary care provider knew they were on this medication: “I don’t want my relationship to be with a bottle of pills and a pamphlet. I want the relationship to be with a trusted PCP …” (#125). Another participant thought that most people would not want to take the pill immediately because they may have questions for their PCP and “… maybe they’ve had more time to think about it as well” (#192).

Most participants did not have strong preferences for the location of follow-up care except to ensure they could get follow-up. Some preferred to follow up with their PCP, assuming they had one they liked, and that the PCP was familiar with PrEP. There was an expectation that the PCP should be able to handle PrEP because “It doesn’t seem like rocket science” (#164). A few participants were concerned about finding a follow-up location after an ED initiation if they did not have a PCP: “If I was to be given like a week of medication and then not be able to get access to care and find a physician, then I just took in those pills for the next five days for no reason” (#064). Similarly, another stated the delay in getting the prescription filled “can cause the potential loss of protection” (#232). Healthcare navigation to connect patients with PrEP-prescribing providers was helpful, with one participant hoping for “… hand holding, … case worker checking in with folks” (#064). Regarding follow-up facilitation, several participants preferred having the PrEP appointments made for them by the ED because of the convenience: “That’d be awesome, one less phone call” (#125).

Additionally, clinical expertise emerged as a priority for participants when considering PrEP follow-up services. One participant receiving care from a provider for university students noted they would prefer to follow up with a physician who had more knowledge about PrEP, “Obviously, I’m going to go to the doctor that has more knowledge in this area” (#192). Similarly, another participant noted the reassurance they would experience seeing an HIV specialist “because if I have any questions, it would be immediately answered on the spot,” and “a specialist has also been exposed to people with similar situations as myself” (#088).

### Key Domain 3: Contextual/ Organizational Factors

#### Benefits and Drawbacks of the ED as a Location for Sexual Health Care

##### More accessible, less stigmatizing

Participants had varying views concerning the provision of PrEP services in the ED. The ED was noted to be a more accessible and less stigmatizing location for addressing sexual health needs than other medical settings. One participant explained that they delayed treatment for syphilis due to fear of seeing a sexual health provider, noting that “…if they’re scared to go to a sexual clinic, they have the option to go to a hospital (i.e., the ED)” (#051).

The ED was also considered a suitable venue for PrEP services because it is where “hard-to- reach” populations presented for care. As one participant explained: “…they’re not seeing their doctor as frequently, or maybe they’re just not educated on public health matters type things so that you will run into a gamut of a variety of people in the ER [ED]” (#064). One participant thought that PrEP in the ED was a “good idea” because “people who go there are already in the mindset of prioritizing their health” (#088). Another noted that education about PrEP could cause a “chain reaction” (#039) of information dissemination, helping to spread awareness among people who might otherwise not receive this information.

There were, however, conflicting thoughts about the appropriateness of receiving preventative and general sexual healthcare in the ED. Three key concerns emerged—the ED’s busyness, the patient’s time burden, and privacy issues.

##### Busyness of the ED

Some participants raised concerns that the ED environment was too busy for specialized PrEP guidance. One worried that ED providers might be too distracted with other tasks to give the highest quality advice: “I just think there’s so much stuff happening that the doctors or the nurses tend to forget to ask if you want to be tested for HIV or pass any other information along…” (#058). Another noted that the urgent and episodic nature of the ED environment may conflict with the prevention mindset needed for PrEP: “I was in a hurry to get out of there” (#125). Similarly, the anticipatory nature of PrEP conflicted with the immediacy of reasons for visiting the ED; one individual stated that PrEP is regarding “what you’re gonna do with the future partners… When you’re in the emergency room it’s because something immediately happened” (#232).

##### Time burden in the ED

When asked if they would spend extra time in the ED to learn about PrEP, participants preferred to minimize extra wait time. However, their willingness to wait depended on how long they had already waited, the emotional stress of the visit, and, as noted above, whether they found PrEP relevant based on their perceived risk. One participant described, “If I’m in this space socially where I think I might need it [PrEP], yes, I would wait about 30 minutes” (#164). Most expressed a willingness to extend their visit up to 20 minutes if it meant receiving valuable information. However, one participant recounted their frustration with feeling unable to leave the ED when they wanted, contributing to their being “anxious to get out of there,” which, in turn, might make them less inclined to stay longer for PrEP information (#125). Similarly, another participant was annoyed with the accumulated time spent in the ED: “I wasted six hours there. I don’t want to stay even one minute there” (#157).

Several participants pointed out that the ED is a convenient location for health promotion activities since patients are already waiting for extended periods: “They’re waiting there for hours, so might as well get additional information…” (#088). One participant noted, “If it’s not making me lose my spot, then I don’t see why not” (#232). This same participant added, “…it just helps kill the time. Also being productive with my health and body.”

##### Privacy concerns

Several participants highlighted privacy concerns related to PrEP services in the ED. Some felt that it would be inappropriate to assess eligibility for PrEP or conduct education in public locations, such as the ED waiting room or with the triage nurse, preferring alternatives like watching a video about PrEP for confidentiality reasons “and things like that of health status” (#051). One questioned the confidentiality of ED procedures based on a previous experience with HIV risk screening: “…there was a patient right next to me, and I believe that that was too close to that [for] these types of questions or even giving me that information” (#164). Discretion was desired for any discussion regarding sexual health, as expressed by one participant: “I don’t want everybody to hear what’s going on with me down there,” because if it was spoken about in a public area, “that might be a little embarrassing” (#039). This participant went on to describe that the triage nurse should not ask about HIV unless it was in a private alcove because “most people are very private.”

## DISCUSSION

In this first qualitative exploration of ED patients’ perspectives on HIV PrEP using the updated CDC 2021 eligibility criteria, we found that participants expressed favorable views of ED-based PrEP services, including screening, education, and initiation of PrEP. They appreciated the opportunity to obtain information they may not have otherwise received about a highly effective medication. They recognized that the ED was a venue where a diverse population of people who could benefit from PrEP were served. Participants also identified important caveats—that PrEP screening and education should be conducted with privacy, that PrEP-related services should not delay other ED care, and that only patients who are not in pain or distress should be offered PrEP services. Additional concerns revolved around contextual factors such as ED busyness—that provider burden could be a barrier to spending time on PrEP services—and where and how they would receive appropriate expertise for PrEP follow-up care. These potential barriers highlight the importance of maintaining privacy within the physical constraints of the ED, integrating PrEP into wait times in the ED workflow, and ensuring linkages to follow-up care as essential items to consider for EDs developing PrEP programs.

Our study extends previous quantitative findings exploring ED-based PrEP as an innovative approach to expand access to and uptake of this important HIV prevention tool.^10,11,20,22–24,32–37^ In a recent review of ED PrEP programs, Gormley et al. found a range in the percentage of PrEP-eligible patients who expressed personal interest in PrEP–from 2%^33^ to 46%^37^ across six studies.^24^ Even higher proportions of PrEP-eligible patients–54%^32^ and 81%^21^, respectively—expressed interest in learning about PrEP in two other studies that measured this outcome.

A notable finding in this study was that several participants indicated they would prefer to start PrEP immediately in the ED rather than receive a pharmacy prescription or referral elsewhere for initiation. The patient’s preference for immediate starts is reflected in prior literature regarding higher rates of PrEP linkage when same-day appointments with PrEP providers were provided during the ED visit,^22^ at an ED-affiliated sexual health clinic,^38^ and a drop-in STI clinic setting.^39^ Linkage is a challenge in every setting where PrEP initiation and/or ongoing PrEP care is not available. Although reviews of PrEP ED programs have found overall low linkage rates to PrEP- initiating sites,^23,24^ more research is needed to understand how these linkage rates may be improved with immediate PrEP appointments or prescriptions.

EDs can play an essential role in increasing PrEP awareness among patients and identifying high-risk individuals who might not be informed about PrEP through other healthcare settings. Participants in this study supported using waiting periods for health promotion and suggested that PrEP education could be integrated into existing downtimes during ED workflows to improve efficiency and engagement. Expanding PrEP services in the ED is supported by the CDC’s 2021 PrEP guidelines, which broadened initiation criteria to engage previously overlooked groups, including heterosexual individuals and cisgender women.^27^

Patient preferences varied regarding whether clinicians or non-clinicians should deliver PrEP education in the ED, highlighting the importance of expertise. Some participants preferred clinicians for their medical training and ability to address complex questions, whereas others valued the approachability and availability of non-clinicians, such as health educators. However, skepticism toward education and screening by peers (described as someone with similar life experiences as you) emerged, with some questioning whether peers’ experiences aligned with their needs and if peers had the expertise to help an individual determine if PrEP was appropriate for them. Participants’ interest in PrEP-knowledgeable providers highlights the need for robust PrEP delivery training for clinical and non-clinical staff to build patient buy-in.

### Limitations

Our findings should be interpreted considering several limitations. The sample was small, representing only 26% of those eligible for the study. Additionally, the findings likely are influenced by social desirability bias, potentially influencing participants’ expressed views to be more favorable to PrEP than reality. Furthermore, this study was conducted at an ED that had not yet formalized PrEP services, and as such, participants’ responses were hypothetical and may not reflect actual behavior if PrEP services were to be offered.

## CONCLUSION

Our study provides insights into patients’ preferences regarding PrEP care in the ED. Key aspects of PrEP preferences include privacy, expertise, and flexible ED workflow integration. These findings can inform the design of patient-centered PrEP programs in emergency care settings.

## Data Availability

The qualitative data supporting the findings of this study are not publicly available due to the sensitive nature of the participant information. However, de-identified excerpts relevant to the key themes discussed in the analysis may be provided upon reasonable request to the corresponding author, with prior approval from the institutional ethics review board

